# Central motor cortical repetitive transcranial magnetic stimulation at 5 Hz or 10 Hz combined with cerebellar 1 Hz stimulation for ataxia in multiple system atrophy of the cerebellar type: a retrospective cohort study of efficacy and predictors of response

**DOI:** 10.64898/2026.11.16.26368149

**Authors:** Wang Yongli, Wang Minghui, Han Yixian, Pan Jiali, Zhou Yongkang, Wang Mengyang

## Abstract

**Objective:** To evaluate the efficacy of central motor cortical repetitive transcranial magnetic stimulation (rTMS) at different frequencies (5 Hz and 10 Hz) combined with cerebellar 1 Hz stimulation on the core domains of ataxia in patients with multiple system atrophy of the cerebellar type (MSA-C), and to explore the independent predictors of the response.

**Methods:** In this retrospective study, 48 patients with a confirmed diagnosis of MSA-C were included. All patients received bilateral cerebellar hemispheric 1 Hz rTMS as background therapy and were divided according to the frequency of central cortical stimulation into a control group (cerebellar 1 Hz stimulation only, n=15), a 5 Hz combination group (n=16) and a 10 Hz combination group (n=17). Ataxia severity before and after treatment was assessed with the International Cooperative Ataxia Rating Scale (ICARS); absolute improvements and improvement rates were calculated for each subscore and for the total score. Continuous variables were described as mean ± standard deviation (SD) or median (interquartile range) according to normality; within-group pre–post comparisons used the Wilcoxon signed-rank test, and between-group comparisons used one-way analysis of variance (ANOVA) or the Kruskal–Wallis H test with Bonferroni correction. Clinical response was defined by an absolute improvement threshold (ΔICARS total score ≥8 points), and independent influencing factors were analysed by Firth-penalised logistic regression; sensitivity analyses were performed with a total-score improvement rate ≥20% (and with ≥15% and ≥30% as alternative thresholds), and robustness results are reported for the ΔICARS ≥10-point criterion. Missing cognitive and mood scale scores were imputed by multiple imputation by chained equations (MICE; m=5; the mean of the five imputations constituted the analysis dataset) and then included in the analysis. Subgroup and interaction analyses were performed according to the median disease duration (2 years) and median age (57 years).

**Results:** After treatment, the ICARS total score in the overall cohort was significantly lower than at baseline (median difference 7.0 points, P<0.001), and posture and gait (ICARS1), limb kinetic function (ICARS2) and speech function (ICARS3) all improved significantly (P<0.001 for all), whereas oculomotor disorders (ICARS4) did not improve significantly (P=0.799). Efficacy differed significantly among the three groups: median total-score improvement rates were 10.7%, 20.3% and 21.9%, respectively (P=0.004); when a total-score improvement rate ≥20% was used to define clinically significant efficacy, the response rates of the three groups were 6.7%, 50.0% and 58.8% (P=0.006), and both the 10 Hz and 5 Hz groups were significantly better than the control group (Bonferroni-corrected P=0.006 and P=0.040), with no statistically significant difference between the two frequencies. Domain-specific comparison showed statistically significant between-group differences in both the absolute improvement and the improvement rate of limb kinetic function (P=0.013 and P=0.005), with the 10 Hz group significantly better than the control group. When clinical response was defined as ΔICARS ≥8 points, the response rates of the three groups were 20.0%, 56.2% and 64.7% (P=0.029); Firth-penalised multivariable regression showed that baseline ICARS total score was an independent predictor of clinical response (OR=1.076, 95% CI 1.013–1.143, P=0.017), whereas 5 Hz (OR=4.298, 95% CI 0.757–24.412, P=0.100) and 10 Hz (OR=5.847, 95% CI 0.885–38.640, P=0.067) stimulation did not reach independent statistical significance after adjustment. When response was defined as ΔICARS ≥10 points, the response rates were 0%, 43.8% and 47.1% (P=0.007), and there were no responders in the control group. Subgroup analysis showed a significant group×age interaction (P=0.020); the clinical response rate of patients aged ≥57 years receiving 10 Hz stimulation was 85.7% (exploratory finding; small subgroup sample). The group×duration interaction was not statistically significant (P=0.210).

**Conclusions:** Central 5 Hz and 10 Hz rTMS combined with cerebellar 1 Hz stimulation both significantly improved the symptoms of ataxia in patients with MSA-C, and the efficacy was domain-specific, with the greatest improvement in the limb kinetic domain; sensitivity analyses across multiple criteria (ΔICARS ≥8 points, improvement rate ≥20%, ΔICARS ≥10 points) consistently supported the efficacy benefit of high-frequency stimulation in the same direction, but stimulation frequency did not reach independent predictive statistical significance after adjustment, and baseline ICARS total score was significantly associated with absolute improvement. Age may have a modifying effect on the efficacy of stimulation frequency (exploratory finding; small sample). This study provides preliminary support for the efficacy of the central 10 Hz combined with cerebellar 1 Hz regimen; the optimal stimulation parameters and independent predictors remain to be verified in larger prospective studies.

## 1. Introduction

Multiple system atrophy (MSA) is a sporadic, rapidly progressive and fatal adult-onset neurodegenerative disease characterised clinically by autonomic dysfunction combined with parkinsonism or cerebellar ataxia, and pathologically by widespread deposition of α-synuclein-containing cytoplasmic inclusions in oligodendrocytes [1,2]. According to the motor phenotype, MSA is divided into a parkinsonian type (MSA-P) and a cerebellar type (MSA-C); MSA-C is characterised by progressive cerebellar ataxia and is relatively more common in Asian populations, accounting for approximately two-thirds of MSA cases [3,4]. Patients with MSA-C usually develop marked gait disturbance and loss of balance within 5 years after onset, resulting in a substantial disease burden [5]; however, no effective disease-modifying therapy able to delay disease progression is yet available, and the options for symptomatic management are limited [6–8].

Repetitive transcranial magnetic stimulation (rTMS) is a non-invasive neuromodulation technique based on the principle of electromagnetic induction that can modulate the excitability of the target cortex and induce synaptic plasticity through repeated pulse delivery; it has shown promising applications in the treatment of a variety of neuropsychiatric diseases [9–11]. Previous studies have confirmed that the neuromodulatory effects of rTMS are frequency-dependent: low-frequency (≤1 Hz) stimulation generally reduces cortical excitability, whereas high-frequency (≥5 Hz) stimulation enhances cortical excitability and can induce more durable plastic changes [12–14]. The cerebello-thalamo-cortical circuit is a core pathway for motor coordination and gait control, and the cortical motor areas (the primary motor cortex M1, the supplementary motor area and others) are both key nodes of this circuit and targets that can be directly modulated by rTMS [15]. In recent years, rTMS studies of cerebellar ataxia have gradually increased: low-frequency cerebellar stimulation can improve truncal ataxia and gait in patients with spinocerebellar degeneration [16,17], and high-frequency or 1 Hz cerebellar stimulation has also produced definite symptomatic improvement in patients with spinocerebellar ataxia (SCA) [18]; small-sample randomised controlled studies in patients with MSA also suggest that cerebellar rTMS can improve ataxia and is accompanied by changes in cortical plasticity [19,20]. Systematic reviews and meta-analyses further support an overall effect of TMS on cerebellar ataxia [21], and the clinical application prospects of non-invasive cerebellar stimulation in neurodegenerative ataxia have also been supported by several reviews [31–33].

However, previous studies have mostly focused on stimulation of a single cerebellar target, and systematic evidence is still lacking on whether there is a frequency–dose-response gradient for the efficacy of central cortical stimulation at different frequencies combined with cerebellar stimulation in MSA-C ataxia, whether different symptom domains respond consistently to treatment, and what the independent predictors of efficacy are. On this basis, the present study retrospectively analysed the efficacy of three regimens — no central stimulation (cerebellar 1 Hz stimulation only), 5 Hz and 10 Hz — each combined with background cerebellar 1 Hz stimulation in the treatment of MSA-C, in order to provide a basis for the selection of individualised clinical treatment parameters.

## 2. Materials and methods

### 2.1. Study participants

Inclusion criteria: (1) patients with probable or possible MSA-C who met the second consensus statement on the diagnosis of multiple system atrophy (2008) or the Movement Disorder Society (MDS) criteria for the diagnosis of multiple system atrophy (2022); (2) patients treated in our hospital between January 2021 and December 2025 with bilateral cerebellar hemispheric 1 Hz rTMS as background therapy (with or without central high-frequency stimulation) who completed efficacy assessments before and after treatment; (3) patients who completed ICARS and cognitive and mood scale assessments before and after treatment. Exclusion criteria: (1) concomitant epilepsy or other diseases of the central nervous system; (2) intracranial metallic foreign bodies or implanted electronic devices (such as cardiac pacemakers); (3) pregnant or lactating women; (4) severe cardiac, pulmonary, hepatic or renal dysfunction or malignancy that would make treatment difficult to tolerate; (5) incomplete core clinical data.

### 2.2. Group allocation and stimulation protocol

All participants received bilateral cerebellar hemispheric 1 Hz rTMS as background therapy. On this basis, participants were divided into three groups according to the frequency of central cortical stimulation: (1) control group (n=15), bilateral cerebellar hemispheric 1 Hz rTMS only, without central stimulation; (2) 5 Hz combination group (n=16), central 5 Hz rTMS combined with cerebellar 1 Hz rTMS; (3) 10 Hz combination group (n=17), central 10 Hz rTMS combined with cerebellar 1 Hz rTMS. Group allocation was based on clinical treatment decisions (retrospective cohort, not randomised allocation).

A neuronavigation-guided transcranial magnetic stimulation system was used to localise the bilateral hand and foot representation areas of the central region and the bilateral cerebellar hemispheres as stimulation targets; the initial stimulation intensity was 80% of the resting motor threshold (RMT) and was subsequently adjusted according to patient tolerance. The stimulation sites included C3, C4 and Cz and the bilateral cerebellar hemispheres (C3, C4 and Cz are the central electrode positions of the 10–20 International System, corresponding to the hand and foot representation areas of the central region localised by neuronavigation). Each site received 800–1000 pulses per session (total course 10 days; total pulses per site 8000–10000), with an interval of 5 min between sites; each session lasted 10–30 min, once daily, for a total course of 10 days. The control group received cerebellar 1 Hz stimulation only, without central stimulation. Apart from the central stimulation frequency (none/5 Hz/10 Hz), stimulation intensity, target localisation, the number of pulses per site and the treatment course were identical among the three groups. Adverse reactions were monitored during treatment in accordance with the safety guidelines for transcranial magnetic stimulation [23]. Concomitant medications (if any) were maintained on the original regimen during treatment and were not adjusted because of stimulation therapy.

### 2.3. Outcome assessment

Ataxia severity was assessed with the International Cooperative Ataxia Rating Scale (ICARS) [22]. The ICARS is a 100-point scale comprising four domains: posture and gait disturbance (0–34 points), limb kinetic function (0–52 points), speech function (0–8 points) and oculomotor disorders (0–6 points), with higher scores indicating more severe impairment. Assessments were performed before treatment and at the end of treatment. Efficacy was rated by neurologists who had received standardised training on the basis of medical records; because the data were derived from routine clinical diagnosis and treatment records, the raters were not blinded to group allocation.

The primary efficacy outcomes were the changes in the ICARS total score and domain scores from before to after treatment (absolute improvement and improvement rate), and the secondary efficacy outcome was the clinical response rate. To facilitate clinical interpretation and further stratification and prediction analyses, a total-score improvement rate ≥20% was pre-specified in this study to define clinically significant efficacy (responders); this cut-off refers to the criterion used in previous ataxia treatment studies in which a ≥20% improvement rate was used to define responders [30], and sensitivity analyses were performed with cut-offs of ≥15% and ≥30%, respectively. In addition, to eliminate the measurement bias that may be introduced by using the baseline score as the denominator when calculating the improvement rate, an absolute improvement threshold was also used to define clinical response and served as the primary criterion for the regression analyses. In the early protocol of this study, a ΔICARS total score ≥10 points was pre-specified to define clinical response; because there were no responders in the control group under this criterion (0/15), which resulted in complete separation and a non-estimable effect size, the primary analysis was changed to a ΔICARS total score ≥8 points (post hoc adjustment), and ΔICARS ≥10 points and a total-score improvement rate ≥20% were used as the robustness and sensitivity criteria, respectively. The results under each criterion are presented side by side and are consistent in direction; no single threshold is regarded as a definitive conclusion in this study.

### 2.4. Statistical analysis

Statistical analysis was performed with Python 3.12 (scipy, statsmodels). Continuous variables were first tested for normality with the Shapiro–Wilk test: variables conforming to a normal distribution were described as mean ± SD (x□±s) and compared among the three groups by one-way analysis of variance (ANOVA); variables not conforming to a normal distribution were described as median (interquartile range) [median (Q1, Q3)] and compared among the three groups by the Kruskal–Wallis H test. Categorical variables were expressed as counts (proportions) and compared between groups by the χ² test or Fisher’s exact test. Within-group pre–post comparisons used the Wilcoxon signed-rank test. Between-group efficacy comparisons used the ICARS total-score improvement rate (continuous variable) and the absolute improvement as the main indicators, the clinically significant response rate (total-score improvement rate ≥20%) as the indicator for stratification and prediction analyses, and sensitivity analyses were performed for response thresholds (≥15%, ≥30%); three-group comparisons used the Kruskal–Wallis H test or ANOVA, pairwise comparisons used the Mann–Whitney U test and the χ²/Fisher exact test, and Bonferroni correction was applied (adjusted significance level α=0.05/3=0.0167). With achievement of clinical response (ΔICARS total score ≥8 points) as the dependent variable, univariable and multivariable Firth-penalised logistic regression analyses were performed to identify independent influencing factors, with group allocation and variables with P<0.20 in univariable analysis entered into the multivariable model; a sensitivity regression was performed using the total-score improvement rate ≥20% criterion, and robustness results are reported for the ΔICARS ≥10-point criterion. Because very few effective events in the control group caused quasi-complete separation of the standard maximum-likelihood estimates, Firth correction was applied in all regressions to obtain stable estimates. Missing cognitive and mood scale scores in the control group (MMSE/MoCA in 6 patients; HAMA/HAMD in 1 patient) were imputed by multiple imputation by chained equations (MICE, m=5); the mean of the five imputed values constituted the final analysis dataset, which was then included in the analysis, with imputed values constrained within the feasible range of each scale. Stratified subgroup analyses were performed according to the median disease duration (2 years) and median age (57 years) (duration was divided into ≤2 years and >2 years; age was divided into <57 years and ≥57 years), and the interaction was tested using the product term of group and stratification factor. In addition, a Firth-penalised regression with disease duration as a continuous variable and a group×duration product term was used to test effect modification. The significance level was α=0.05 (two-sided).

## 3. Results

### 3.1. Baseline clinical characteristics

The three groups did not differ significantly in age, sex distribution, disease duration, years of education, baseline ICARS total score or domain scores, HAMA, HAMD or MoCA (all P>0.05). MMSE differed significantly among the three groups (P=0.008), with a lower baseline MMSE score in the control group, and MoCA showed a borderline trend towards a difference (P=0.056). Apart from the cognitive scores, baseline clinical characteristics were generally balanced and comparable; the between-group difference in MMSE was included as a covariate for adjustment in the subsequent regression analyses (see 3.4.1) (Table 1).

**Table 1.** Baseline clinical characteristics of the three groups.

| **Characteristic** | **All patients (n=48)** | **Control (n=15)** | **5 Hz (n=16)** | **10 Hz (n=17)** | **P value** |
|---|---|---|---|---|---|
| Age, years, mean ± SD | 57.5±7.5 | 57.5±6.5 | 58.7±7.7 | 56.5±8.3 | 0.716 |
| Sex, male/female | 25/23 | 8/7 | 8/8 | 9/8 | 0.979 |
| Education, years, median | 12.0 (9.0, 15.0) | 12.0 (9.0, 13.5) | 10.5 (9.0, 12.8) | 12.0 (9.0, 16.0) | 0.497 |
| Disease duration, years, median | 2.0 (2.0, 3.0) | 2.5 (2.0, 3.0) | 2.0 (1.9, 3.0) | 2.0 (2.0, 3.0) | 0.646 |
| ICARS1 posture and gait, mean ± SD | 15.9±6.7 | 15.8±5.4 | 16.6±7.3 | 15.4±7.4 | 0.834 |
| ICARS2 limb kinetic function, mean ± SD | 19.8±5.7 | 20.5±4.5 | 20.4±5.7 | 18.6±6.7 | 0.576 |
| ICARS3 speech function, mean ± SD | 2.7±1.4 | 2.7±1.2 | 2.8±1.6 | 2.5±1.5 | 0.946 |
| ICARS4 oculomotor disorders, mean ± SD | 1.8±1.3 | 1.7±1.6 | 1.9±1.3 | 1.7±1.2 | 0.725 |
| ICARS total score, mean ± SD | 40.1±12.5 | 40.7±10.3 | 41.7±11.6 | 38.2±15.2 | 0.724 |
| HAMA, median | 5.0 (3.0, 9.3) | 5.0 (3.5, 11.5) | 6.0 (3.0, 8.2) | 4.0 (3.0, 9.0) | 0.437 |
| HAMD, median | 10.5 (5.8, 15.3) | 10.0 (6.5, 21.5) | 13.0 (7.8, 15.0) | 6.0 (4.0, 13.0) | 0.111 |
| MMSE, median* | 25.0 (23.0, 27.3) | 23.0 (21.0, 25.0) | 25.0 (22.8, 28.0) | 27.0 (25.0, 28.0) | 0.008 |
| MoCA, median | 22.0 (18.0, 23.3) | 19.0 (16.5, 22.5) | 21.0 (19.2, 23.2) | 23.0 (21.0, 25.0) | 0.056 |

ICARS1–ICARS4 denote the four domains of the International Cooperative Ataxia Rating Scale (ICARS). HAMA, Hamilton Anxiety Rating Scale; HAMD, Hamilton Depression Rating Scale; MMSE, Mini-Mental State Examination; MoCA, Montreal Cognitive Assessment. Normally distributed variables are expressed as mean ± SD and non-normally distributed variables as median (Q1, Q3). \*MMSE differed significantly among the three groups (P=0.008), with a lower baseline MMSE score in the control group; MoCA showed a borderline trend towards a difference (P=0.056).

### 3.2. Changes in ICARS domain scores in the overall cohort

#### 3.2.1. Overall efficacy

After treatment, the ICARS total score in the overall cohort was significantly lower than at baseline, with a median difference of 7.0 (3.8, 10.2) points (Wilcoxon signed-rank test, P<0.001) and a mean improvement rate (including only patients with a baseline score >0) of 19.07%±17.50%, indicating that combined central and cerebellar rTMS has a definite improving effect on ataxia symptoms in patients with MSA-C. Changes in each domain before and after treatment are detailed in Table 2 and Figure 1.

**Figure 1.**
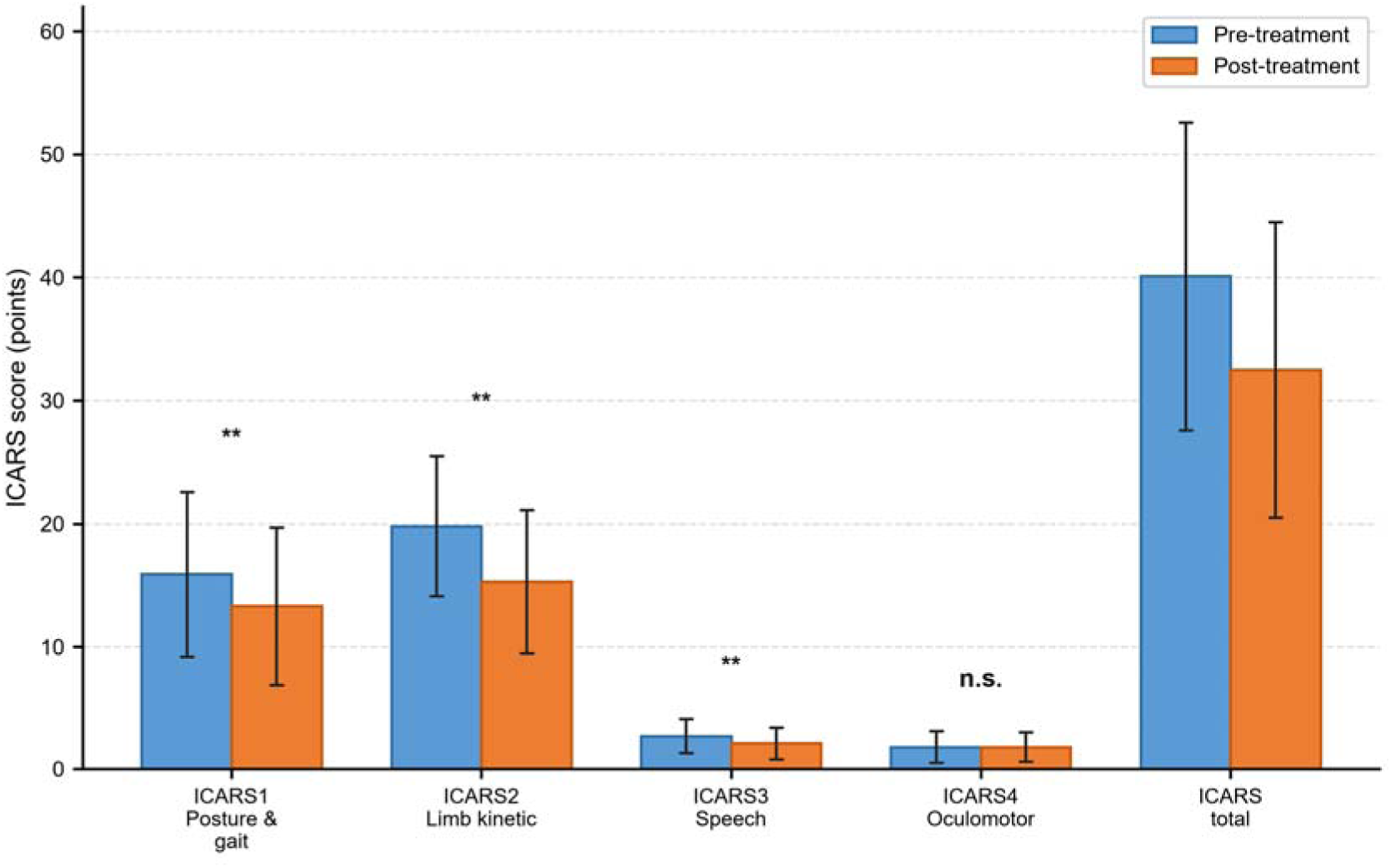
ICARS domain scores before and after treatment in the overall cohort (n=48), showing mean values with standard deviation error bars; **P<0.001 and n.s., not significant (Wilcoxon signed-rank test)

**Table 2.**
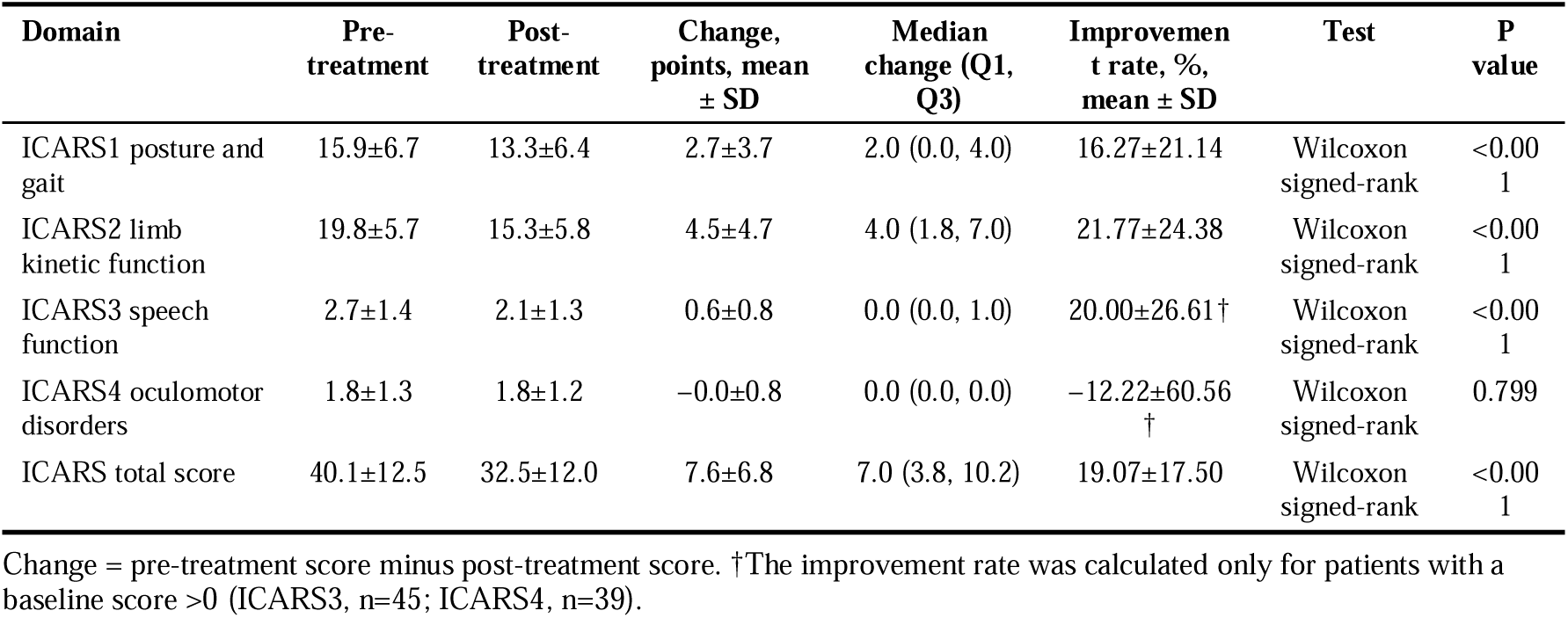
ICARS domain scores before and after treatment in the overall cohort.

#### 3.2.2. Heterogeneity of domain-specific responses

The treatment responses of the four ICARS domains showed some heterogeneity: (1) limb kinetic function (ICARS2) showed the greatest improvement, with a mean reduction of 4.5 ± 4.7 points after treatment and a median difference of 4.0 (1.8, 7.0) points (P<0.001); (2) posture and gait (ICARS1) showed a mean reduction of 2.7 ± 3.7 points, with a median difference of 2.0 (0.0, 4.0) points (P<0.001); (3) speech function (ICARS3) showed a mean reduction of 0.6 ± 0.8 points, a difference that was statistically significant (P<0.001), but the median difference was 0, suggesting that the extent of improvement in speech symptoms was limited in most patients; (4) oculomotor disorders (ICARS4) showed no statistically significant difference in scores before and after treatment (P=0.799), suggesting that the present rTMS regimen has no clear improving effect on oculomotor symptoms.

### 3.3. Differences in efficacy among stimulation frequencies

Baseline ICARS scores were balanced among the three groups, and efficacy after treatment differed between groups (Table 3 and Figure 2). The median total-score improvement rates of the three groups were 10.7% in the control group, 20.3% in the 5 Hz group and 21.9% in the 10 Hz group, with a statistically significant between-group difference (Kruskal–Wallis H=10.883, P=0.004; the Kruskal–Wallis test served as the primary test, and ANOVA F=6.505, P=0.003 is shown as supplementary information). Patients who did not reach the 20% threshold also improved to varying degrees (for example, the median improvement rate in the control group was 10.7%), and this efficacy information is fully represented by the continuous improvement-rate indicator. When a total-score improvement rate ≥20% was used to define clinically significant efficacy, the response rates of the three groups were 6.7% (1/15), 50.0% (8/16) and 58.8% (10/17), with a statistically significant between-group difference (χ²=10.153, P=0.006). When response was defined by absolute improvement, the ΔICARS ≥8-point response rates were 20.0% (3/15), 56.2% (9/16) and 64.7% (11/17), with a statistically significant between-group difference (χ²=7.049, P=0.029); pairwise comparisons did not reach statistical significance after Bonferroni correction (adjusted P=0.086 for 0 vs 10 Hz). The ΔICARS ≥10-point response rates were 0% (0/15), 43.8% (7/16) and 47.1% (8/17), with a significant between-group difference (Fisher’s exact test P=0.007); there were no responders with ΔICARS ≥10 points in the control group, and pairwise comparisons gave adjusted P=0.009 for 0 vs 10 Hz and adjusted P=0.020 for 0 vs 5 Hz. Sensitivity analyses with cut-offs of ≥15% and ≥30% showed between-group differences in the same direction as with the ≥20% threshold (P=0.010 and P=0.021), indicating that the conclusions were robust. Pairwise comparisons (Bonferroni-corrected) showed that both the improvement rate and the response rate in the 10 Hz group were significantly better than in the control group (P=0.006 and P=0.008), and the 5 Hz group was also significantly better than the control group (P=0.040 and P=0.046); the differences between the 5 Hz and 10 Hz groups were not statistically significant (P=1.000).

**Figure 2.**
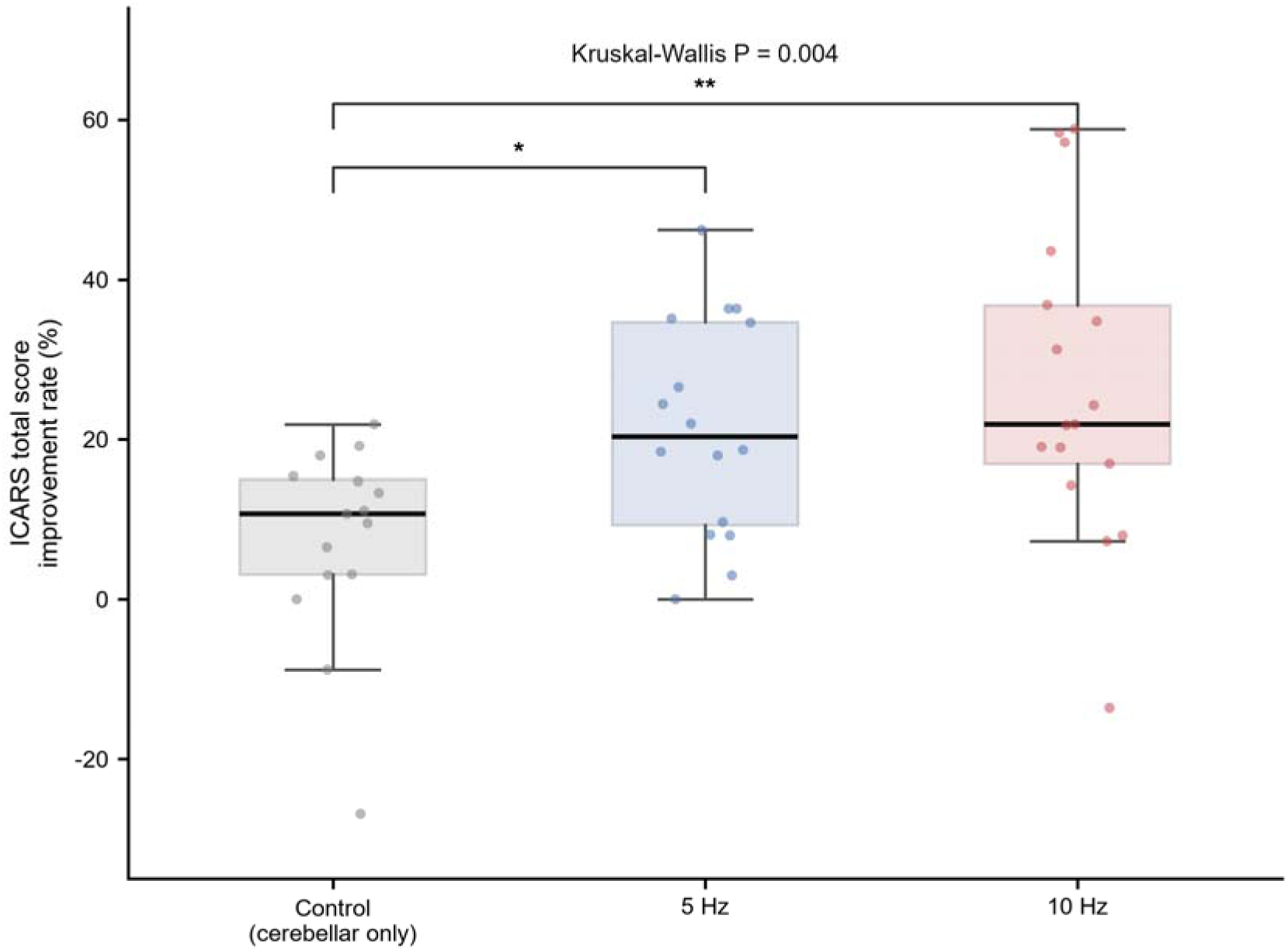
ICARS total-score improvement rate in the three groups (box plots with median and interquartile range and individual values overlaid); Kruskal–Wallis P=0.004, *P=0.040 and **P=0.006 versus the control group (Bonferroni-corrected pairwise comparisons). Control, cerebellar 1 Hz stimulation only

**Table 3.** Efficacy and changes in ICARS domains in the three groups.

| Outcome | Control (n=15) | 5 Hz (n=16) | 10 Hz (n=17) | Test | P value |
|---|---|---|---|---|---|
| ICARS total-score improvement rate, % | 10.7 (3.1, 15.1) | 20.3 (9.3, 34.7) | 21.9 (16.9, 36.8) | Kruskal–Wallis H | 0.004 |
| ΔICARS total score, points | 3.6±4.6 | 8.8±5.7 | 10.1±7.9 | Kruskal–Wallis H | 0.012 |
| Clinically significant response, n (%) | 1/15 (6.7) | 8/16 (50.0) | 10/17 (58.8) | χ<sup>2</sup> test | 0.006 |
| ΔICARS1 posture and gait, points | 1.2±2.2 | 3.3±3.5 | 3.4±4.6 | Kruskal–Wallis H | 0.221 |
| ΔICARS2 limb kinetic function, points‡ | 1.7±3.0 | 4.7±4.3 | 6.6±5.1 | Kruskal–Wallis H | 0.013 |
| ΔICARS3 speech function, points | 0.5±0.6 | 0.8±1.0 | 0.5±0.6 | Kruskal–Wallis H | 0.785 |
| ΔICARS4 oculomotor disorders, points | 0.2±0.9 | 0.1±0.4 | −0.4±0.9 | Kruskal–Wallis H | 0.271 |
| ICARS1 improvement rate, % | 6.2 (0.0, 17.1) | 18.5 (6.2, 33.3) | 18.2 (0.0, 42.9) | Kruskal–Wallis H | 0.169 |
| ICARS2 improvement rate, %‡ | 8.7 (2.2, 17.8) | 17.9 (6.2, 32.5) | 31.6 (18.2, 45.5) | Kruskal–Wallis H | 0.005 |

Δ denotes the pre- to post-treatment difference, expressed as mean ± SD; improvement rates are expressed as median (Q1, Q3) and were calculated only for patients with a baseline score >0. ‡Compared with the control group, Bonferroni-corrected P=0.013 (ΔICARS2) and P=0.005 (ICARS2 improvement rate). Pairwise comparisons (Bonferroni-corrected): total-score improvement rate, 0 vs 5 Hz P=0.040, 0 vs 10 Hz P=0.006, 5 vs 10 Hz P=1.000; clinically significant response rate, 0 vs 5 Hz P=0.046, 0 vs 10 Hz P=0.008, 5 vs 10 Hz P=1.000. Clinically significant efficacy was defined as an ICARS total-score improvement rate ≥20%; in sensitivity analyses with cut-offs of ≥15% (26.7%, 68.8%, 76.5%; P=0.010) and ≥30% (0%, 31.2%, 41.2%; P=0.021), the direction of the between-group differences was consistent. When response was defined by absolute improvement: ΔICARS ≥8-point response rates 20.0%/56.2%/64.7% (χ<sup>2</sup>=7.049, P=0.029); ΔICARS ≥10-point response rates 0%/43.8%/47.1% (Fisher's exact test P=0.007; no responders in the control group); between-group P=0.601 and P=0.440 for ICARS3 and ICARS4 improvement rates (these two rows are not shown).

Domain-specific between-group comparisons showed that the differences in efficacy were domain-specific: for limb kinetic function (ICARS2), the between-group differences in both absolute improvement and improvement rate were statistically significant (P=0.013 and P=0.005), and the 10 Hz group was significantly better than the control group (median absolute improvement 5.0 vs 2.0 points, Bonferroni-corrected P=0.013; median improvement rate 31.6% vs 8.7%, adjusted P=0.005), whereas the differences between the 5 Hz group and the control group did not reach statistical significance (adjusted P=0.228 and P=0.245); for the posture and gait, speech and oculomotor domains, between-group differences in absolute improvement were not statistically significant (P=0.221, P=0.785 and P=0.271), and the improvement rates of the speech and oculomotor domains also did not differ statistically between groups. These findings suggest that the differences in efficacy among frequencies are mainly expressed in the limb kinetic domain.

### 3.4. Analysis of independent influencing factors

#### 3.4.1. Logistic regression analysis of clinical response

With a ΔICARS total score ≥8 points defining clinical response (the primary analysis criterion), the response rates of the three groups and the results of pairwise comparisons are given in section 3.3 and the footnote to Table 3. The results of Firth-penalised univariable and multivariable logistic regression are shown in Table 4 and Figure 3: after multivariable adjustment, neither 10 Hz (OR=5.847, 95% CI 0.885–38.640, P=0.067) nor 5 Hz (OR=4.298, 95% CI 0.757–24.412, P=0.100) stimulation reached independent predictive statistical significance; baseline ICARS total score was an independent predictor of clinical response (OR=1.076, 95% CI 1.013–1.143, P=0.017), suggesting that patients with more severe baseline ataxia symptoms were more likely to reach the threshold of absolute improvement. A sensitivity regression with the total-score improvement rate ≥20% as the dependent variable showed significant positive associations for 5 Hz (OR=35.167, 95% CI 1.691–731.381, P=0.021) and 10 Hz (OR=58.833, 95% CI 1.943–1781.579, P=0.019), but the ORs and confidence intervals were extreme, and because the dependent variable contains the baseline score as a denominator the estimates are affected by measurement coupling; these results are therefore provided for robustness reference only. When response was defined as ΔICARS ≥10 points, there were no responders in the control group (0/15), and no finite OR could be estimated because of complete separation (response rates and pairwise comparisons are given in section 3.3). The results under the various criteria were consistent in direction and support an improvement in efficacy with high-frequency stimulation, but the conclusion that stimulation frequency is an independent predictor was not supported under the absolute-improvement criterion and requires verification in larger samples.

**Figure 3.**
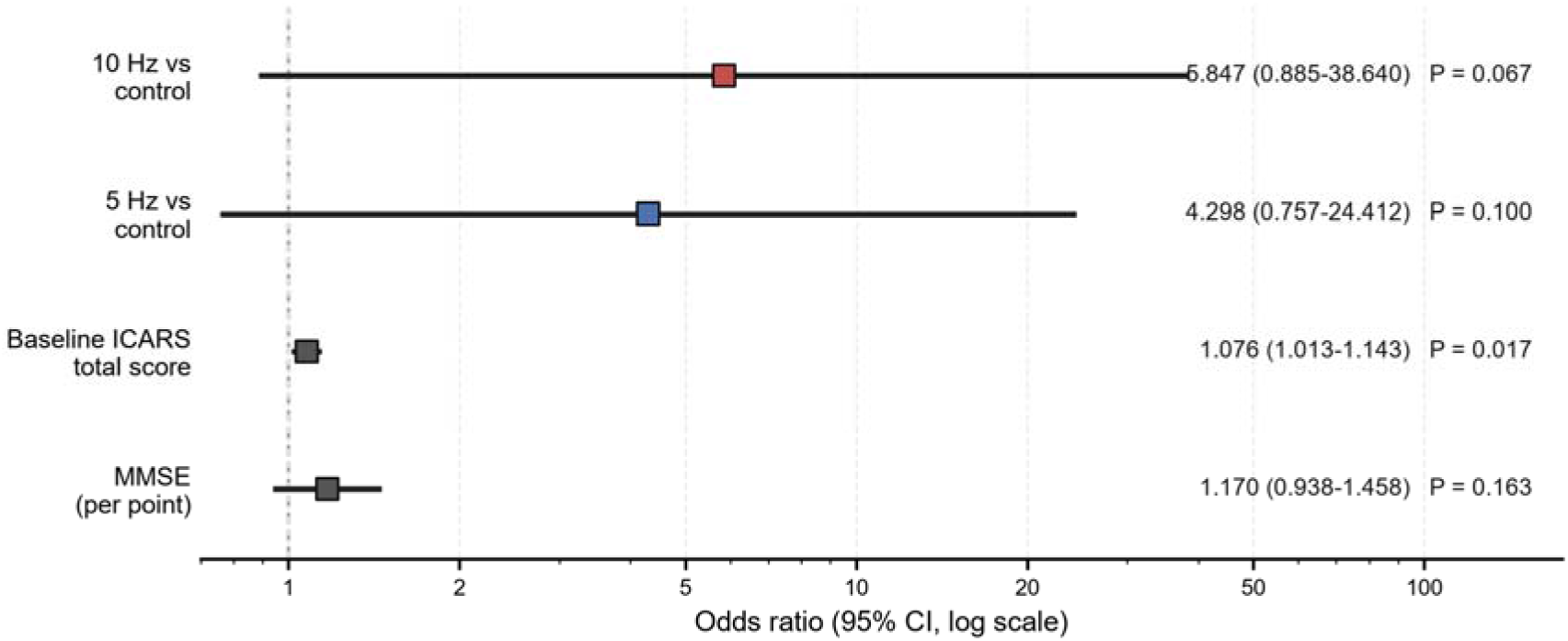
Multivariable Firth-penalised logistic regression for clinical response defined by absolute improvement (ΔICARS ≥8 points) (model M1, log scale; the size of the squares is proportional to the effect size and horizontal lines indicate 95% confidence intervals)

**Table 4.** Univariable and multivariable Firth-penalised logistic regression for clinical response defined by absolute improvement (ΔICARS ≥8 points)

| Variable | Univariable OR (95% CI) | P value | Multivariable OR (95% CI) | P value |
|---|---|---|---|---|
| 5 Hz vs control | 1.616 (0.482–5.413) | 0.436 | 4.298 (0.757–24.412) | 0.100 |
| 10 Hz vs control | 2.760 (0.811–9.393) | 0.104 | 5.847 (0.885–38.640) | 0.067 |
| Age | 0.953 (0.880–1.033) | 0.243 | — | — |
| Sex (male) | 0.730 (0.234–2.274) | 0.587 | — | — |
| Disease duration | 1.416 (0.823–2.437) | 0.209 | — | — |
| Years of education | 1.003 (0.880–1.143) | 0.966 | — | — |
| Baseline ICARS total score | 1.058 (1.004–1.114) | 0.036 | 1.076 (1.013–1.143) | 0.017 |
| HAMA | 1.043 (0.926–1.175) | 0.483 | — | — |
| HAMD | 1.037 (0.961–1.118) | 0.347 | — | — |
| MMSE | 1.201 (0.994–1.451) | 0.058 | 1.170 (0.938–1.458) | 0.163 |
| MoCA | 1.063 (0.937–1.206) | 0.343 | — | — |

Clinical response was defined as a ΔICARS total score ≥8 points; OR, odds ratio; CI, confidence interval; the control group (cerebellar stimulation only) served as the reference. Univariable analysis included all candidate variables (Firth-penalised); the multivariable model (Firth-penalised, M1) included group allocation and variables with P<0.20 in univariable analysis (baseline ICARS total score, MMSE), and because very few effective events in the control group caused quasi-complete separation of the standard maximum-likelihood estimates, Firth correction was applied in all regressions. Sensitivity analyses: with the total-score improvement rate ≥20% as the dependent variable, 5 Hz OR=35.167 (1.691–731.381), P=0.021 and 10 Hz OR=58.833 (1.943–1781.579), P=0.019; interpretation is limited because the dependent variable contains the baseline score as a denominator and because the confidence intervals are extreme. With ΔICARS ≥10 points as the dependent variable, 0/15 responders in the control group led to complete separation and the OR was not estimable (response rates 0%, 43.8% and 47.1%; Fisher's exact test P=0.007). The direction of results was consistent across criteria.

### 3.5. Subgroup analyses

The results are shown in Table 5 and Figures 4 and 5.

**Figure 4.**
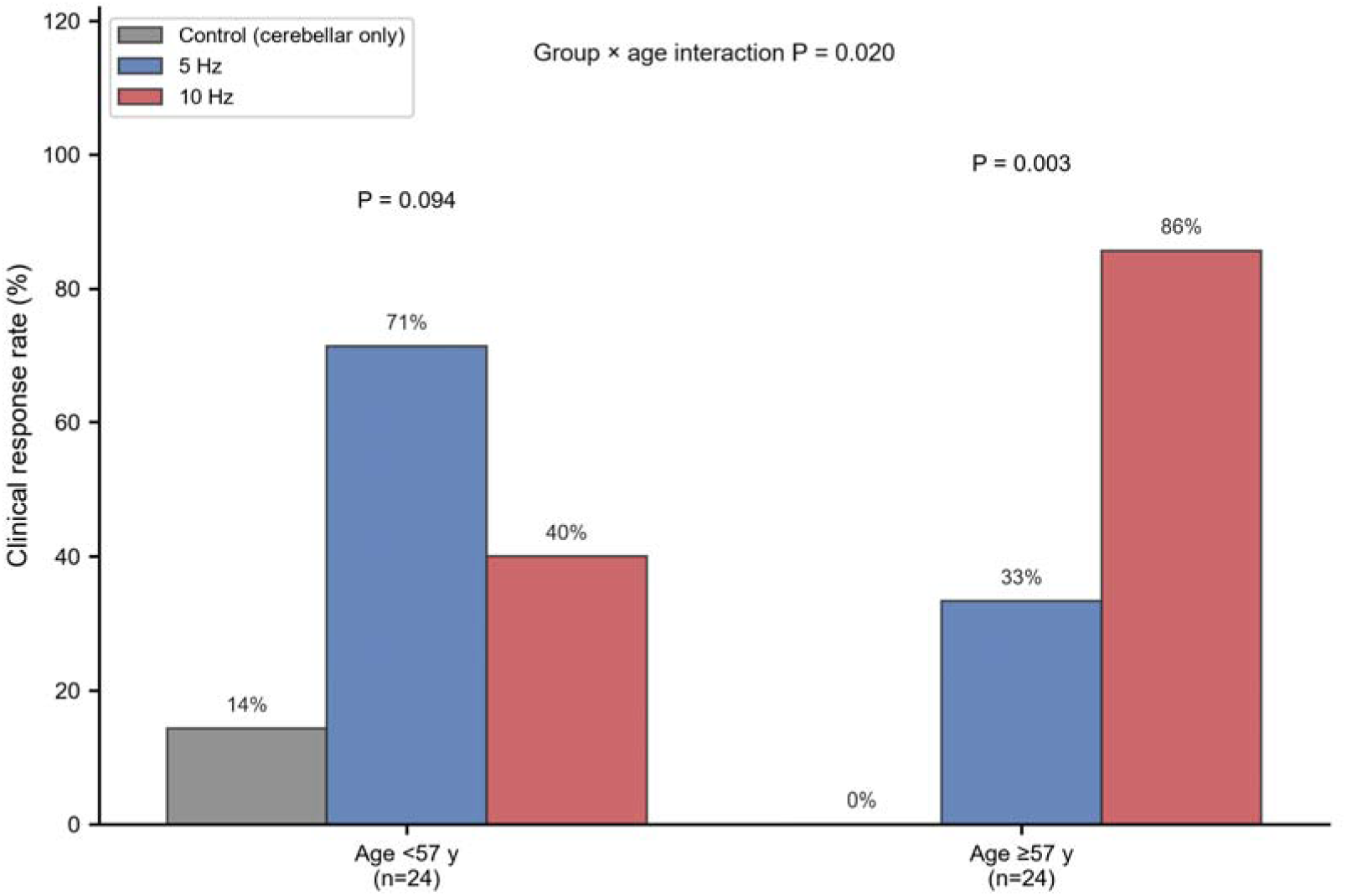
Clinically significant response rate by age stratum in the three groups. Clinical response was defined as an ICARS total-score improvement rate ≥20%; the group×age interaction P value was 0.020

**Figure 5.**
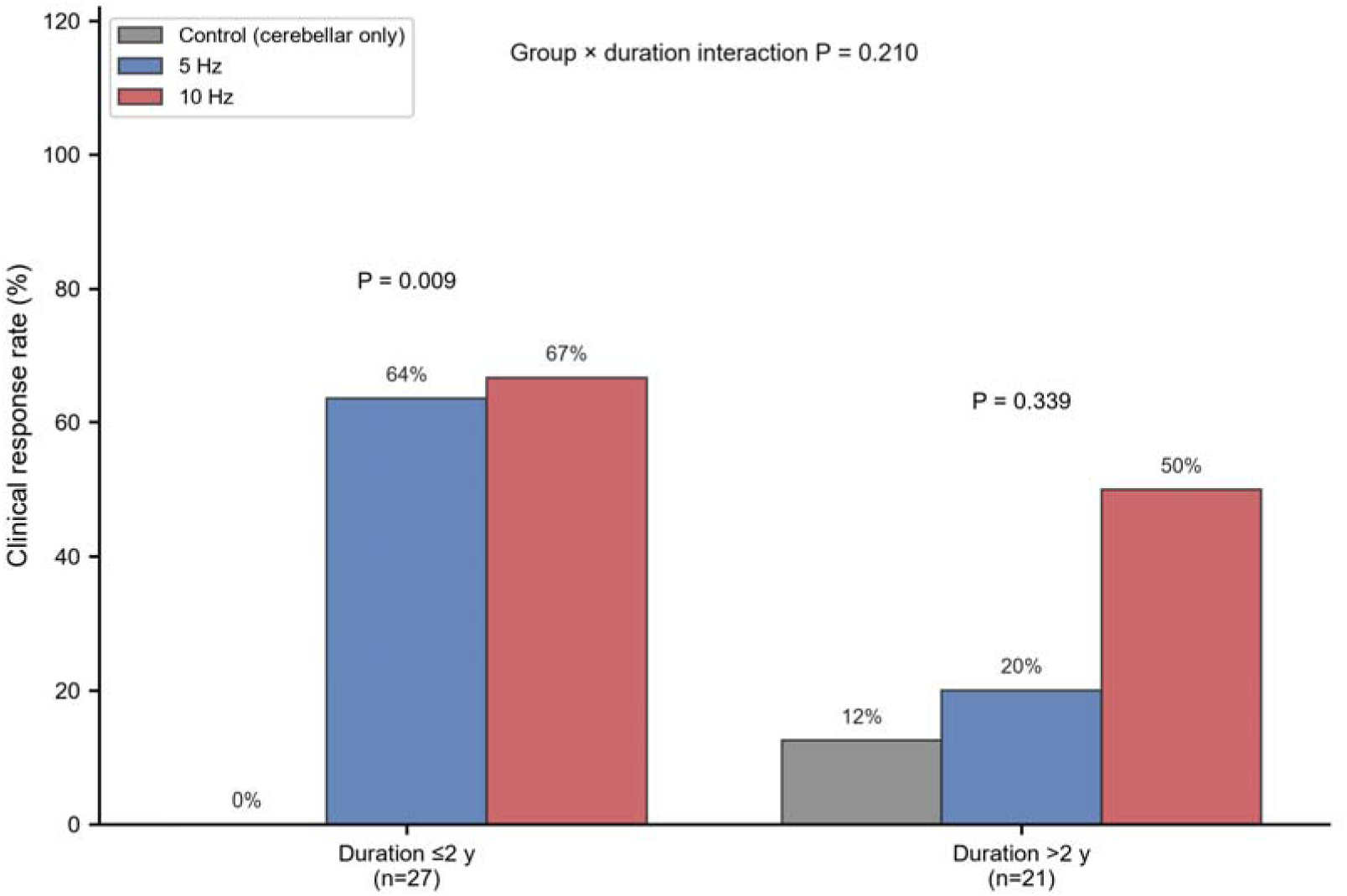
Clinically significant response rate by disease duration stratum in the three groups. Clinical response was defined as an ICARS total-score improvement rate ≥20%; the group×duration interaction P value was 0.210

**Table 5.**
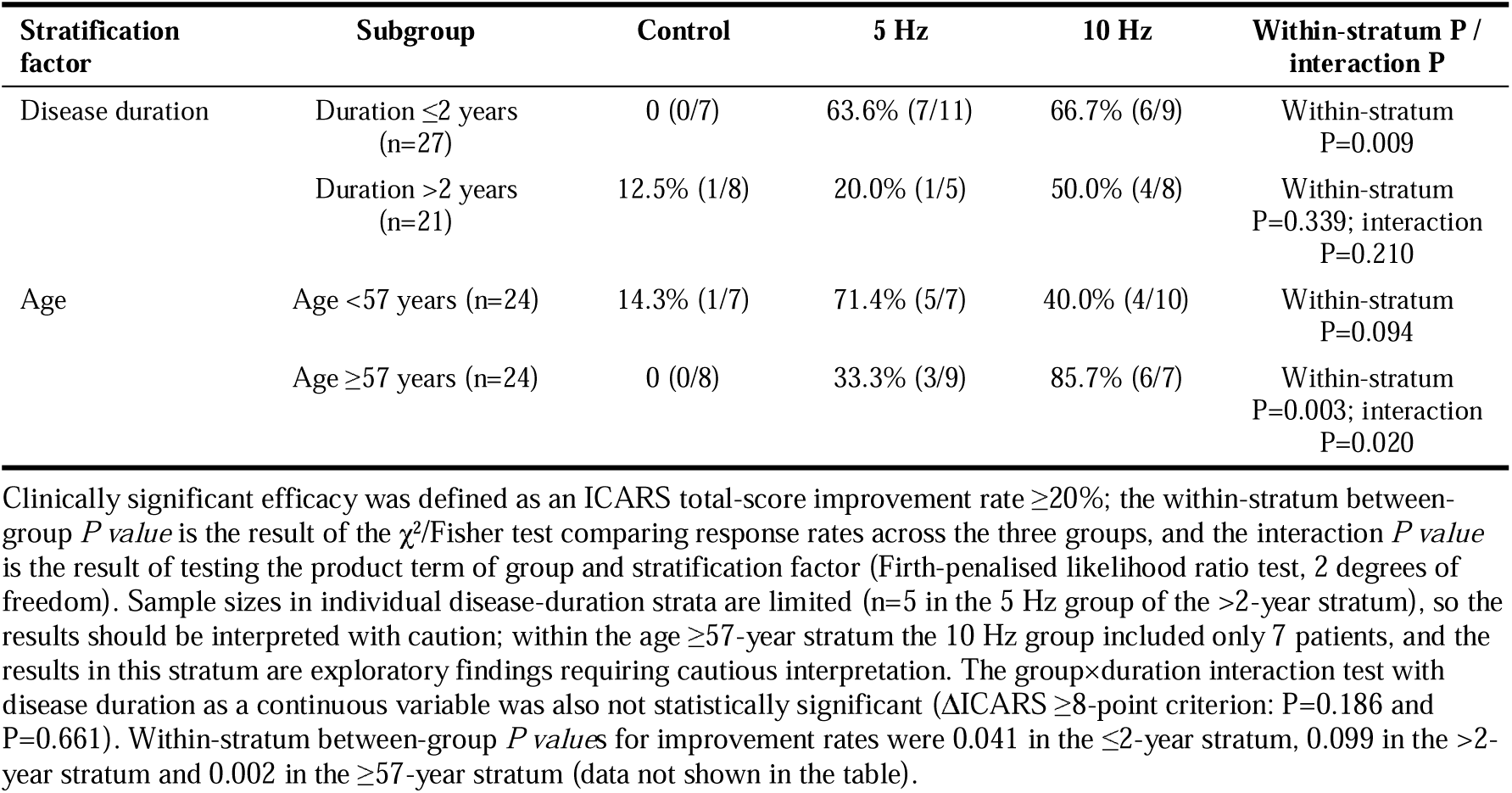
Clinically significant response rates in different subgroups.

| **Stratification factor** | **Subgroup** | **Control** | **5 Hz** | **10 Hz** | **Within-stratum P / interaction P** |
|---|---|---|---|---|---|
| Disease duration | Duration ≤2 years (n=27) | 0 (0/7) | 63.6% (7/11) | 66.7% (6/9) | Within-stratum P=0.009 |
|  | Duration >2 years (n=21) | 12.5% (1/8) | 20.0% (1/5) | 50.0% (4/8) | Within-stratum P=0.339; interaction P=0.210 |
| Age | Age <57 years (n=24) | 14.3% (1/7) | 71.4% (5/7) | 40.0% (4/10) | Within-stratum P=0.094 |
|  | Age ≥57 years (n=24) | 0 (0/8) | 33.3% (3/9) | 85.7% (6/7) | Within-stratum P=0.003; interaction P=0.020 |

Clinically significant efficacy was defined as an ICARS total-score improvement rate ≥20%; the within-stratum between-group *P value* is the result of the χ²/Fisher test comparing response rates across the three groups, and the interaction *P value* is the result of testing the product term of group and stratification factor (Firth-penalised likelihood ratio test, 2 degrees of freedom). Sample sizes in individual disease-duration strata are limited (n=5 in the 5 Hz group of the >2-year stratum), so the results should be interpreted with caution; within the age ≥57-year stratum the 10 Hz group included only 7 patients, and the results in this stratum are exploratory findings requiring cautious interpretation. The group×duration interaction test with disease duration as a continuous variable was also not statistically significant (ΔICARS ≥8-point criterion: P=0.186 and P=0.661). Within-stratum between-group *P value*s for improvement rates were 0.041 in the ≤2-year stratum, 0.099 in the >2-year stratum and 0.002 in the ≥57-year stratum (data not shown in the table).

Disease-duration stratification: within the ≤2-year stratum (n=27), the clinically significant response rates of the three groups differed statistically significantly (0% in the control group, 63.6% in the 5 Hz group and 66.7% in the 10 Hz group; P=0.009), and both the 5 Hz and 10 Hz groups were significantly higher than the control group; within the >2-year stratum (n=21), the between-group difference did not reach statistical significance (12.5%, 20.0% and 50.0%; P=0.339), although the response rate in the 10 Hz group remained numerically higher. The group×duration interaction was not statistically significant (P=0.210). Firth regression interaction tests with disease duration as a continuous variable and a group×duration product term also did not reach statistical significance (ΔICARS ≥8-point criterion: 5 Hz×duration P=0.186, 10 Hz×duration P=0.661; the interactions were also non-significant under the improvement rate ≥20% criterion and after multivariable adjustment), suggesting that the modifying effect of disease duration on the efficacy of stimulation frequency was not significant in this study and that the conclusion was robust across multiple cut-offs, criteria and the continuous-variable framework.

Age stratification: within the ≥57-year stratum (n=24), both the improvement rates (P=0.002) and the clinically significant response rates (P=0.003) of the three groups differed statistically significantly, and the clinically significant response rate in the 10 Hz group was as high as 85.7% (6/7), significantly higher than in the 5 Hz group (33.3%) and the control group (0%); the 10 Hz group in this subgroup included only 7 patients, so the results are exploratory findings requiring cautious interpretation; within the <57-year stratum the difference did not reach statistical significance (P=0.094). The group×age interaction was statistically significant (P=0.020), suggesting that age has a significant modifying effect on the efficacy of stimulation frequency and that patients aged ≥57 years benefit particularly prominently from 10 Hz stimulation.

### 3.6. Safety

All patients tolerated treatment well during the treatment period; no serious adverse events (such as induced seizures or syncope) were observed, and no patient discontinued treatment or withdrew from the study because of adverse reactions.

## 4. Discussion

This study systematically evaluated the differences in efficacy of central rTMS at different frequencies combined with cerebellar 1 Hz stimulation across the core domains of ataxia in patients with MSA-C, and analysed the independent predictors of efficacy. The study found that the improvement of MSA-C ataxia by rTMS was domain-specific: the limb kinetic domain showed the greatest improvement and a significant frequency-related difference in efficacy, and the efficacy advantage of 10 Hz and 5 Hz high-frequency stimulation was clear, with sensitivity analyses across multiple criteria consistently supporting an efficacy benefit in the same direction; however, after adjustment for confounders under the absolute-improvement criterion (ΔICARS ≥8 points), the independent predictive effect of stimulation frequency did not reach statistical significance. Age had a significant modifying effect on the efficacy of stimulation frequency, and patients aged ≥57 years benefited particularly prominently from 10 Hz stimulation; the modifying effect of disease duration did not reach statistical significance.

### 4.1. Mechanisms underlying the domain specificity of rTMS for MSA-C ataxia

In this study, among the four ICARS domains, posture and gait (ICARS1), limb kinetic function (ICARS2) and speech function (ICARS3) all improved significantly compared with before treatment, with the greatest improvement in limb kinetic function (median difference 4.0 points), whereas oculomotor disorders (ICARS4) showed no clear benefit. Between-group comparisons showed that the differences in efficacy among frequencies were mainly expressed in the limb kinetic domain, in which both the absolute improvement and the improvement rate of the 10 Hz group were significantly better than those of the control group, with the 5 Hz group intermediate between the two.

The maintenance of posture and gait and the coordination of limb movements both depend on the concerted action of the cerebello-thalamo-cortical circuit, the spinocerebellar tracts and subcortical motor networks, in which cortical structures such as the primary motor cortex and the supplementary motor area are direct targets of rTMS modulation [15,24]. Central rTMS can reshape the impaired cerebello-cortical circuit by modulating cortical excitability, thereby improving motor planning and execution and consequently alleviating limb ataxia and gait disturbance. Previous studies have shown that rTMS applied to the primary motor cortex or the cerebellar region can significantly improve gait stability and balance in patients with cerebellar ataxia [16,18,20], consistent with the domain-specific results of this study. Limb kinetic function mainly reflects the fine coordination of limb movements, and its associated neural circuits are closely connected with the cortical motor areas, making it particularly sensitive to high-frequency central stimulation; this accords with the significant frequency-related difference in efficacy observed in this domain in the present study. In contrast, speech dysfunction and oculomotor disorders are more closely related to involvement of deep structures such as the brainstem cranial nerve nuclei, the cerebellar vermis and the medial longitudinal fasciculus in the brainstem, and the induced electric field of transcranial magnetic stimulation can hardly reach the deep nuclei effectively [15]; the improving effect is therefore limited. In addition, oculomotor disorders showed a numerically slight worsening trend in the 10 Hz group (not statistically significant); whether this is related to stimulation remains unclear, and the mechanism requires further investigation.

### 4.2. Dose–response relationship of stimulation frequency and its clinical implications

The results of this study showed that, when a total-score improvement rate ≥20% was used to define clinically significant efficacy, the response rates of central 5 Hz and 10 Hz stimulation combined with cerebellar 1 Hz (50.0% and 58.8%, respectively) were both significantly higher than that of the control group (6.7%), whereas the difference between the two frequencies was not statistically significant; in the multivariable analysis under the absolute-improvement criterion (ΔICARS ≥8 points), neither 10 Hz (OR=5.847, 95% CI 0.885–38.640) nor 5 Hz (OR=4.298, 95% CI 0.757–24.412) stimulation reached independent statistical significance, and the 95% confidence intervals of the ORs for the two frequencies were wide and overlapped; the present data therefore cannot support the existence of a discernible dose–response gradient between frequencies, and verification in larger samples is required. This pattern is consistent with the basic electrophysiological evidence for rTMS — low-frequency stimulation generally inhibits cortical excitability, whereas high-frequency stimulation can more effectively induce cortical plastic changes and produce more sustained neuromodulatory effects [12–14] — and also with the results of rTMS studies in neurological diseases such as Parkinson’s disease and post-stroke motor impairment [26]. At present, the level of evidence-based recommendation for cerebellar ataxia in international rTMS guidelines is still accumulating [10,11], and this study provides direct clinical evidence for the selection of a regimen of central high-frequency stimulation combined with cerebellar stimulation. It should be pointed out that clinically significant efficacy (total-score improvement rate ≥20%) served only as an indicator for efficacy stratification and prediction analysis in this study, and that the main body of the efficacy evidence is the between-group comparison of continuous improvement rates; patients who did not reach this threshold also improved to varying degrees (for example, the median improvement rate in the control group was 10.7%), and this efficacy information is fully retained through the continuous indicator. Sensitivity analyses (≥15%, ≥30%) showed that the direction of between-group differences was consistent across thresholds, indicating that the conclusions are robust.

Considering that patients with MSA-C are mostly middle-aged and older adults who often have comorbidities such as cerebrovascular disease and epilepsy, the tolerability of high-frequency stimulation may differ between individuals [23]; in clinical practice, the stimulation frequency can therefore be selected individually according to the patient’s tolerance. It is noteworthy that recent studies suggest that patterned stimulation (such as iTBS and cTBS) also has the potential to modulate ataxia when applied to the cerebellar region [19,25,27], and the optimal combination of different stimulation modes and frequencies is an important direction for future research.

### 4.3. Clinical implications of the predictors of efficacy

This study found that, under the absolute-improvement threshold criterion (ΔICARS ≥8 points), stimulation frequency did not reach independent predictive statistical significance after adjustment for confounders (10 Hz OR=5.847, P=0.067; 5 Hz OR=4.298, P=0.100), whereas baseline ICARS total score was an independent predictor of clinical response (OR=1.076, P=0.017), suggesting that patients with more severe baseline symptoms were more likely to reach the threshold of absolute improvement; when the total-score improvement rate ≥20% was used as the dependent variable, frequency showed significant positive associations (ORs 35.167 and 58.833, respectively), but these estimates are affected by the improvement rate–baseline coupling and have extreme confidence intervals, which limits interpretation. The results under the various criteria consistently support an improvement in efficacy with high-frequency stimulation, but the conclusion that frequency is an independent predictor was not supported under the absolute criterion and requires verification in larger prospective studies. Subgroup analyses suggest that age may have a modifying effect on the efficacy of stimulation frequency: patients aged ≥57 years had a higher response rate with 10 Hz stimulation (85.7%) than the 5 Hz group and the control group, whereas no significant between-frequency difference was observed in patients aged <57 years. This suggests that older patients with MSA-C may have a stronger neuromodulatory response to high-frequency central stimulation, but the sample size of this subgroup is small (7 patients in the 10 Hz group) and the finding is exploratory; the specific mechanism requires further clarification in combination with neurophysiological studies. In the disease-duration subgroups, patients with a duration ≤2 years (relatively early disease) had markedly higher clinically significant response rates with high-frequency stimulation (63.6% in the 5 Hz group and 66.7% in the 10 Hz group vs 0% in the control group; within-stratum P=0.009), consistent with the general principle in progressive neurodegenerative diseases that patients with a shorter disease course retain better neuroplasticity and benefit more from early intervention [1,7,8]; in patients with a duration >2 years, the between-group difference did not reach statistical significance (P=0.339), although the response rate in the 10 Hz group remained numerically higher (50.0%). The interaction of disease duration did not reach statistical significance (interaction P=0.210 for the ≤2/>2-year stratification; P=0.186 and P=0.661 for the group×duration interaction test with duration as a continuous variable; P=0.156 for the stratified interaction under the ΔICARS ≥8-point criterion), suggesting that the modifying effect of disease duration on efficacy is not supported by sufficient statistical evidence, which may be related to the sample size of this study.

It is noteworthy that, under the ΔICARS ≥8-point criterion, univariable analysis showed a borderline trend for the baseline MMSE score (OR=1.201, P=0.058), which did not reach statistical significance after multivariable adjustment (P=0.163); moreover, the baseline MMSE score of the control group was lower (P=0.008), and this imbalance in the cognitive baseline may have a potential confounding influence on the comparison of efficacy, so the relevant results should be interpreted with caution. Sex, years of education and disease duration had no independent predictive effect on clinically significant efficacy, suggesting that this treatment regimen has good generalisability across the population and that patients need not be excluded from treatment on account of demographic characteristics.

### 4.4. Limitations

This study has several limitations. First, it was a single-centre retrospective analysis with a relatively small sample (48 patients) and limited statistical power; participants were allocated on the basis of clinical treatment decisions rather than randomly, so selection bias is possible, and because clinical response events in the control group were few, the confidence intervals of the ORs after Firth correction were wide (for example, 95% CI 0.885–38.640 for 10 Hz), so the results should be interpreted with caution and require further verification in large-sample multicentre studies.

Second, the control group received no central stimulation (bilateral cerebellar 1 Hz stimulation only) and no complete sham stimulation control was included, so placebo effects and the contribution of the intrinsic efficacy of cerebellar stimulation to the results cannot be completely excluded; in addition, efficacy was assessed on the basis of routine medical records and the raters were not blinded, so assessment bias cannot be completely excluded.

Third, the baseline MMSE score of the control group was lower (P=0.008), and this imbalance in the cognitive baseline may have a confounding influence on the comparison of efficacy; concomitant medications were also not analysed statistically in this study, so their potential confounding effects could not be completely excluded.

Fourth, only the short-term efficacy after treatment was assessed and no long-term follow-up was performed, so the disease-modifying effect of rTMS on the progression of MSA-C remains unclear.

Fifth, this study explored only the effects of 5 Hz, 10 Hz or no central stimulation combined with cerebellar 1 Hz; the differences in efficacy of modes such as intermittent theta burst stimulation (iTBS) and continuous theta burst stimulation (cTBS) still require further investigation [27,29].

Sixth, no neurophysiological (such as TMS-EEG) or imaging indices were included, so the evidence for the mechanisms of cortical plasticity after stimulation is limited [19,21].

Seventh, the definition of clinical response in this study involved several reasonable criteria (among which the primary analysis criterion of ΔICARS ≥8 points was a post hoc adjustment made after the pre-specified ΔICARS ≥10-point criterion could not be estimated because of complete separation): under the absolute-improvement threshold, stimulation frequency did not reach independent predictive significance whereas baseline ICARS total score was independently significant; under the improvement rate ≥20% criterion, frequency was significant but the estimate was affected by dependent variable–baseline measurement coupling; under the ΔICARS ≥10-point criterion, there were no responders in the control group, resulting in complete separation. The results across criteria were consistent in direction but the effect estimates were unstable, and the related conclusions require verification in larger samples. Missing cognitive and mood scale data in the control group were imputed by MICE (m=5) to constitute the analysis dataset; the uncertainty introduced by imputation itself was not completely eliminated, and no separate complete-case sensitivity analysis was performed.

## 5. Conclusions

Central 5 Hz and 10 Hz rTMS combined with cerebellar 1 Hz stimulation both significantly improved the symptoms of ataxia in patients with MSA-C, and the efficacy was domain-specific: the posture and gait, limb kinetic function and speech function domains all improved significantly, with the greatest improvement in the limb kinetic domain and no clear benefit for oculomotor disorders; the differences in efficacy among frequencies were mainly expressed in the limb kinetic domain.

Both 5 Hz and 10 Hz central stimulation combined with cerebellar 1 Hz significantly improved MSA-C ataxia; when absolute improvement (ΔICARS ≥8 points) was used as the response criterion, stimulation frequency did not reach independent predictive statistical significance after adjustment, and baseline ICARS total score was significantly associated with absolute improvement; sensitivity analyses under the improvement rate ≥20% and ΔICARS ≥10-point criteria were consistent in direction and support the efficacy benefit of high-frequency stimulation, but the effect estimates are limited, and confirmation of frequency as an independent predictor requires larger samples. Age had an exploratory modifying effect on the efficacy of stimulation frequency, with the highest response rate in patients aged ≥57 years receiving 10 Hz stimulation.

These results provide preliminary support for the efficacy of central 10 Hz combined with cerebellar 1 Hz stimulation in improving MSA-C ataxia, particularly for limb kinetic impairment; its long-term efficacy, the optimal stimulation parameters and the independent predictors remain to be further verified in large-sample, double-blind, sham-controlled prospective studies.

## Data Availability

All data produced in the present study are available upon reasonable request to the authors
All data produced in the present work are contained in the manuscript

## Abbreviations

ANOVA: analysis of variance
cTBS: continuous theta burst stimulation
CI: confidence interval
HAMA: Hamilton Anxiety Rating Scale
HAMD: Hamilton Depression Rating Scale
ICARS: International Cooperative Ataxia Rating Scale
iTBS: intermittent theta burst stimulation
MDS: Movement Disorder Society
MICE: multiple imputation by chained equations
MMSE: Mini-Mental State Examination
MoCA: Montreal Cognitive Assessment
MSA: multiple system atrophy
MSA-C: multiple system atrophy of the cerebellar type
MSA-P: multiple system atrophy of the parkinsonian type
OR: odds ratio
RMT: resting motor threshold
rTMS: repetitive transcranial magnetic stimulation
SCA: spinocerebellar ataxia
SD: standard deviation
TMS-EEG: transcranial magnetic stimulation combined with electroencephalography.

## Declarations

### Ethics approval and consent to participate

This study was approved by the Ethics Committee of Sanbo Brain Hospital, Capital Medical University (approval no. SBNK-YJ-202009-01). As the study was a retrospective analysis of routine clinical records, the requirement for written informed consent was waived by the committee. All procedures were performed in accordance with the Declaration of Helsinki.

### Consent for publication

Not applicable.

### Availability of data and materials

The de-identified data analysed in this study are available from the corresponding author on reasonable request.

### Authors’ contributions

Wang.M.H. and Han.Y.X. conceived and designed the study (Conceptualization, Methodology). Pan.J.L. and Zhou.Y.K. delivered the repetitive transcranial magnetic stimulation and collected the clinical data (Investigation, Resources). Wang.Y.L. and Wang.M.H. analysed the clinical data and performed the statistical analysis (Formal Analysis, Validation). Wang.M.Y.contributed to the interpretation and visualization of the data (Visualization, Writing – Review & Editing). Wang.Y.L drafted the manuscript (Writing – Original Draft). Wang.M.Y. supervised the project and acquired funding (Supervision, Funding Acquisition). All authors read and approved the final manuscript and agree to be accountable for all aspects of the work.

## Acknowledgements

We thank the patients whose routine clinical data were analysed in this study, and their families. We are grateful to the clinical, nursing and technical staff of Sanbo Brain Hospital, Capital Medical University, for their support in delivering repetitive transcranial magnetic stimulation and in completing the pre- and post-treatment assessments.

## Registration

Not applicable (retrospective cohort study).

